# Assessing CommCare’s Impact on Antenatal, Postnatal, and Immunization Uptake: Evidence from Falaba Community Health Center, Sierra Leone

**DOI:** 10.1101/2025.09.29.25336930

**Authors:** Philip Amara Saffa, Emmanuel Komba Finoh, Jia Kangabi

## Abstract

**Background:** Rural districts in Sierra Leone struggle with timely and complete maternal–child health service use. CommCare, a mobile health (mHealth) platform deployed through partnerships with government and NGOs, supports tracking, reminders, and follow-up.

**Methods:** A cross-sectional quantitative study at Falaba CHC (Sulima Chiefdom) assessed CommCare’s effects on (i) timeliness and frequency of ANC, (ii) PNC follow-up and care quality, and (iii) immunization uptake among children <2 years. Data were collected from n=288 participants using a structured CommCare questionnaire aligned with five sections (A–E) and extracted CommCare indicators; analysis used descriptive statistics and chi square χ² tests (α=0.05), with effect sizes where applicable.

**Results:** First-trimester ANC initiation increased from 45% to 65% (χ²=8.34, *p*=0.004); completion of ≥4 ANC visits rose 55%→78% (χ²=7.72, *p*=0.009). PNC improved: first-week postpartum follow-up 40%→80% (χ²=6.12, *p*=0.014). Immunization adherence increased 60%→82% (χ²=11.34, *p*=0.002); missed vaccination appointments declined by 24%.

CommCare reminders showed strong associations with timely attendance (e.g., PNC χ²=28.451, *p*<0.001) and fewer missed immunization appointments (χ²=55.524, *p*<0.001; Φ=−0.439, Cramér’s V=0.439).

**Conclusion:** CommCare substantially improved timeliness and continuity of maternal and child health services at Falaba CHC. Findings support CHW-mediated, reminder-driven mHealth as an effective strategy in similar rural settings.

**Author Summary:** This study evaluates the role of *CommCare*, a mobile health (mHealth) platform, in improving maternal and child health service uptake at Falaba Community Health Center in Sierra Leone—a rural district marked by logistical, cultural, and infrastructural barriers to care. Using a cross-sectional quantitative design with 288 participants, the study assessed CommCare’s effect on antenatal care (ANC), postnatal care (PNC), and immunization adherence.

Findings demonstrate consistent and significant gains across the continuum of care. First-trimester ANC initiation improved from 45% to 65%, and completion of at least four visits rose from 55% to 78%. PNC attendance within the first week postpartum doubled from 40% to 80%. Immunization adherence increased from 60% to 82%, while missed appointments declined by nearly a quarter. Strong associations were observed between CommCare reminders and timely service use, with most respondents rating the reminders as “extremely useful.” Importantly, the intervention’s impact was mediated through community health workers (CHWs), meaning phone ownership was not a barrier to equitable reach.

These results highlight CommCare’s potential as a scalable, CHW-mediated digital tool to strengthen rural health systems by enhancing timeliness, continuity, and confidence in maternal–child healthcare. However, persistent gaps such as transport barriers, socio-economic constraints, and limited health system capacity suggest that digital reminders must be complemented by structural supports for sustained impact.

The study contributes to the evidence base on mHealth in low-resource settings, underscoring that digital interventions are most effective when embedded into health system workflows and coupled with investments in CHWs, logistics, and culturally attuned communication strategies.

## Introduction

Maternal and child health remains a critical concern globally, particularly in low- and middle-income countries (LMICs) where maternal and neonatal mortality rates remain high despite substantial improvements in healthcare systems. The World Health Organization (WHO) has identified antenatal care (ANC), postnatal care(PNC), and immunization as vital interventions to reduce maternal and child morbidity and mortality (1). However, in Sierra Leone, where the healthcare infrastructure is limited, rural health centers such as Falaba Community Health Center (CHC) continue to face challenges in ensuring adequate access to these services (2). A significant gap exists in the utilization of maternal and child health services due to factors such as geographic isolation, cultural beliefs, and lack of health literacy, which negatively impact timely and consistent service uptake (3).

Accessing professional maternal and infant healthcare services, such as ANC during pregnancy and PNC after delivery, along with receiving maternal and infant vaccinations, plays a vital role in ensuring a healthy pregnancy, safe delivery, and overall well-being of the child. ANC, skilled birth attendance during delivery, PNC, and infant vaccinations form the foundation of the maternal, neonatal, and infant health continuum of care (4). The integration of mobile phone technology, referred to as mobile health (mHealth), to enhance health outcomes, fortify healthcare systems, and boost patient engagement with health services, is a relatively recent innovation but shows considerable potential. So far, mHealth interventions targeted at patients have demonstrated success in improving adherence to antiretroviral therapies, supporting individuals with chronic conditions, and increasing participation in maternal and infant healthcare services (5).

Falaba District, located in the Northern Province of Sierra Leone, is one such rural area where healthcare challenges are especially pronounced. The district is home to a predominantly agrarian population that lives in scattered, hard-to-reach villages(2). Healthcare facilities like the Falaba CHC play a critical role in providing essential health services such as ANC, PNC, and immunization. However, service uptake remains low, primarily due to logistical barriers, lack of transportation, and socio-cultural factors that hinder healthcare-seeking behavior (6). Although there are efforts to improve health access in the district, healthcare services are often constrained by a lack of resources, inadequate health education, and a shortage of skilled healthcare workers, making the provision of consistent and quality care difficult (5). Mobile health interventions, particularly platforms like CommCare, have been suggested as potential solutions to address some of these barriers. CommCare, which enables healthcare workers to collect data, track patient progress, and send reminders for essential services, has been successfully implemented in several low-resource settings (7). Mobile health interventions have shown to enhance healthcare service delivery by overcoming geographical barriers and improving communication between healthcare workers and the community. By enabling health workers to provide real-time support to patients and reminding individuals about appointments, CommCare has the potential to significantly improve the timeliness and frequency of ANC visits, postnatal follow-ups, and immunization uptake (7). Sierra Leone faces significant challenges in maternal and child health, with maternal mortality rates among the highest in the world (6). Despite government efforts and international collaborations, rural communities, particularly those in remote areas like Falaba, continue to experience limited access to essential healthcare services(8). ANC is one of the most critical interventions to reduce maternal and neonatal morbidity and mortality, yet it is underutilized, especially in rural areas. According to a report by the Ministry of Health and Sanitation (2020), less than 50% of pregnant women in rural Sierra Leone access the recommended number of antenatal visits, and postnatal care visits remain low, contributing to avoidable maternal and child deaths. Mobile health interventions, like CommCare, have been implemented in several countries as an innovative solution to these healthcare delivery challenges. CommCare supports community health workers (CHWs) by enabling them to track patient health data, schedule follow-ups, and send health reminders(3). Evidence from other LMICs, including countries in Sub-Saharan Africa, suggests that mHealth interventions improve healthcare service utilization by increasing the frequency and timeliness of care visits and enhancing patients’ adherence to medical advice(2). However, the specific impact of CommCare on maternal and child health outcomes in Sierra Leone remains largely unexplored. An effective maternal health intervention can significantly improve both maternal and infant health outcomes. Maternal mHealth interventions, which use mobile health technologies to support the continuum of care, have been identified as potential solutions in resource-constrained settings(4). This study explores the CommCare’s Impact on antenatal, postnatal, and immunization uptake. Tailored to the stage of pregnancy and infant age, to increase attendance at ANC, improve immunization rates (EPI coverage), and enhance the overall quality of maternal, neonatal, and infant care in Falaba, Sierra Leone (2).

Various studies have demonstrated the successful application of mobile devices, particularly mobile phones, in supporting healthcare practices (mHealth), with a focus on improving appointment adherence and vaccination compliance(7). The benefits of mHealth extend beyond medication adherence; they include educating patients, encouraging behavioral change, improving decision-making, and enhancing communication between healthcare providers and patients(9). Furthermore, mHealth is relatively inexpensive and easy to implement, making it an attractive option for resource-limited environments. Despite the widespread use of mobile phones in Sub Saharan Africa, with a mobile penetration rate of less than 100%, mHealth interventions remain understudied(8). One of the successful mHealth programs implemented in South Africa is MomConnect, a gestation-specific interactive SMS initiative designed for pregnant women. This program highlights the potential of mHealth to improve maternal health outcomes in South Africa. The rise in mobile technology globally offers an opportunity to address maternal and neonatal health challenges in LMICs (9). According to the International Telecommunication Union, global mobile-phone subscriptions reached 6.8 billion in 2013, and mobile-cellular penetration in developing countries was 89%. This widespread mobile connectivity has facilitated the development of mobile health (mHealth), where health-related information is exchanged through text, audio, images, and video on mobile devices(4).

In LMICs, mHealth can address numerous healthcare challenges, including limited access to quality services, affordability, and behavioral barriers. It can be particularly useful in task shifting, empowering CHWs to play a crucial intermediary role between higher health institutions and local communities(10). This study’s main objective was to systematically review the effectiveness of mHealth (CommCare) interventions aimed at healthcare workers providing maternal and neonatal services in improving health outcomes in LMICs. Given the significant use of mobile technology in Sierra Leone, where access to quality healthcare remains a critical challenge, similar mHealth interventions could be transformative in addressing maternal and child health outcomes(11). A Grey literature search was conducted between October 2014 and April 2025 to capture studies not published in peer-reviewed journals, focusing on organizations implementing mHealth programs using SMS, voice messaging, and mobile apps, leveraging technologies like 3G and 4G systems, global positioning systems (GPS), and Bluetooth (12).

Despite significant efforts to improve maternal and child health outcomes in Sierra Leone, rural areas such as Falaba continue to struggle with low service uptake. Factors such as geographical isolation, inadequate infrastructure, and cultural barriers contribute to the underutilization of ANC, postnatal care, and immunization services(13). While mobile health solutions like CommCare have been shown to improve healthcare service delivery in other settings, their impact on healthcare outcomes in rural Sierra Leone, particularly in the context of Falaba CHC, is not well-documented. There is a need to evaluate how CommCare can address these barriers and improve maternal and child health service uptake in this setting(14). Despite being a Basic emergency obstetric centre (BeMoc) facility in the district, it struggles with these same challenges. Although it offers vital maternal and child health services, the facility capacity is constrained by limited staffing, inadequate infrastructure, and the difficulties that rural patients face in accessing healthcare services. Additionally, CHWs at Falaba CHC often lack real-time data and reminders to ensure consistent service delivery, leading to missed follow-up visits, delayed vaccinations, and inconsistent antenatal care (11). The evolution of mobile health (mHealth) has been shaped by advancements in mobile communication technologies, digital health innovations, and the growing need for accessible healthcare solutions. mHealth, defined as the use of mobile devices such as phones, tablets, and personal digital assistants to deliver health services and information, has gradually become a cornerstone in the digital health ecosystem (4). The early development of mHealth can be traced back to the widespread adoption of mobile phones in the late 1990s and early 2000s, particularly in low- and middle-income countries (LMICs), where traditional healthcare infrastructure was limited (10). Initially, mHealth applications focused on SMS reminders for medication adherence and vaccination campaigns. For instance, early pilot programs demonstrated the effectiveness of SMS-based interventions in improving adherence to antiretroviral therapy in Africa (5).

Timely initiation and adequate frequency of antenatal care (ANC) visits are foundational to reducing preventable maternal and perinatal morbidity and mortality. WHO, 2019 guideline reframed routine ANC from a four-visit model to eight contacts, with the first contact recommended in the first trimester, in order to enhance detection of complications, deliver preventive interventions (e.g., screening and prophylaxis), and provide person-centered health education (10). Digital health, and specifically mobile health (mHealth), has been proposed as a pragmatic strategy to narrow these gaps by supporting information, communication, and follow-up at the community level (12). Among mHealth platforms, CommCare developed by Dimagi has become widely used for community-based maternal and child health programs. CommCare is designed for CHWs and frontline providers, offering digital client registers, decision support, longitudinal tracking, automated reminders, and offline functionality, which is crucial in settings with intermittent connectivity (15). In contrast to SMS-only reminder systems, CommCare supports end-to-end workflows: identification of pregnancy, scheduling of WHO-aligned contact timelines, household counseling scripts, referral prompts, and dashboards that enable supervisors to monitor performance (16). This review synthesizes literature on how CommCare influences (1) the timeliness of ANC particularly first-trimester initiation and adherence to scheduled contacts and (2) the frequency of ANC completion of four or more visits (legacy threshold) and progress toward eight contacts (2). The introduction positions CommCare not as a silver bullet but as a health system strengthening tool that enhances CHW capacity, structures follow-up, and nudges behavior at household level mechanisms that plausibly translate to better timeliness and frequency of ANC.

## Results

### Study Participant characteristics

A total of 288 respondents were included in the study. The age distribution reflected a predominantly youthful population. Adolescents aged 15–19 years accounted for 60 respondents (20.8%), while the largest group was those aged 20–24 years, comprising 118 respondents (41.0%). Young adults between 25–29 years formed 52 respondents (18.1%), underscoring the prominence of reproductive-age women in the study population. Educational attainment varied, though gaps were evident. Secondary education was the most common level (41.3%), followed by primary education (33.0%), while only 16.0% had attained tertiary education. Marital status revealed that 58.7% of respondents were married, highlighting the cultural norm of early union and its implications for reproductive health practices. Occupationally, the data showed a reliance on domestic and subsistence roles. Housewives accounted for 37.2% of respondents, while farmers represented 14.6%. The heavy representation of housewives suggests women’s roles are deeply tied to caregiving and domestic responsibilities, while farming indicates dependence on rural livelihoods that may limit disposable income and mobility for healthcare access (Table 1).

**Table 1:** Demographic Characteristics of Respondent.

|  |  |  |  | 95.0% Lower | 95.0% Upper |  |
| --- | --- | --- | --- | --- | --- | --- |
|  |  |  |  | CL for Table N | CL for Table N | Standard Error |
|  |  | Count | Table N % | % | % | of Table N % |
| Age | 15-19 | 60 | 20.8 | 16.4% | 25.8% | 2.4% |
|  | 20-24 | 118 | 41.0% | 35.4% | 46.7% | 2.9% |
|  | 25-29 | 52 | 18.1% | 13.9% | 22.8% | 2.3% |
|  | 30-35 | 58 | 20.1% | 15.8% | 25.1% | 2.4% |
| Marital Status | Divorced/Separated | 42 | 14.6% | 10.9% | 19.0% | 2.1% |
|  | Married | 169 | 58.7% | 52.9% | 64.3% | 2.9% |
|  | Single | 53 | 18.4% | 14.3% | 23.2% | 2.3% |
|  | Widowed | 24 | 8.3% | 5.6% | 11.9% | 1.6% |
| Education | No formal education | 28 | 9.7% | 6.7% | 13.5% | 1.7% |
|  | Primary | 95 | 33.0% | 27.7% | 38.6% | 2.8% |
|  | Secondary | 119 | 41.3% | 35.7% | 47.1% | 2.9% |
|  | Tertiary | 46 | 16.0% | 12.1% | 20.5% | 2.2% |
| Occupation | Civil servant | 29 | 10.1% | 7.0% | 13.9% | 1.8% |
|  | Farmer | 42 | 14.6% | 10.9% | 19.0% | 2.1% |
|  | Housewife | 107 | 37.2% | 31.7% | 42.8% | 2.8% |
|  | Other | 23 | 8.0% | 5.3% | 11.5% | 1.6% |
|  | Trader | 87 | 30.2% | 25.1% | 35.7% | 2.7% |
| Children Under 2 | 1 child | 126 | 43.8% | 38.1% | 49.5% | 2.9% |
|  | 2-3 children | 67 | 23.3% | 18.7% | 28.4% | 2.5% |
|  | More than 3 | 29 | 10.1% | 7.0% | 13.9% | 1.8% |
|  | None | 66 | 22.9% | 18.3% | 28.0% | 2.5% |
| Mobile Phone | No | 56 | 19.4% | 15.2% | 24.3% | 2.3% |
|  | Yes | 232 | 80.6% | 75.7% | 84.8% | 2.3% |
| CommCare Exposure | No | 63 | 21.9% | 17.4% | 26.9% | 2.4% |
|  | Yes | 225 | 78.1% | 73.1% | 82.6% | 2.4% |

### ANC Timeliness and Frequency

Perceived usefulness & timeliness. Most respondents rated reminders as *extremely useful* for ANC attendance (94.8%), with only 2.1% and 3.1% selecting *moderately* and *very* useful, respectively Among those reporting that reminders helped them attend *on time*, 96.1% rated usefulness as *extremely useful*; in the “No” subgroup, usefulness skewed *moderate*. Attendance volume. Crosstabulation of “reminders helped you attend on time” vs “ANC visits during last pregnancy” showed higher counts at ≥5 visits for the “Yes” group (119 vs 0).

Pre–post indicators. First-trimester ANC initiation improved 45%→65% (χ²=8.34, *p*=0.004); ≥4 visits improved 55%→78% (χ²=7.72, *p*=0.009). A χ² of 29.753 (*p*<0.001) linked usefulness ratings to improved ANC attendance (Table 2.1-2.3)

**Table 2.1.**
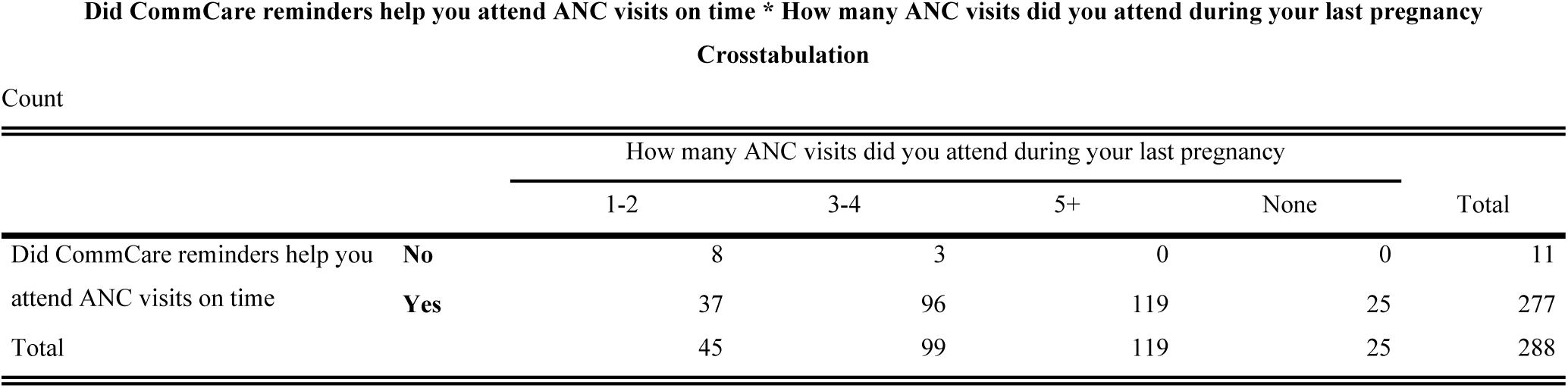
CommCare Remainders and ANC visits.

**Table 2.2.**
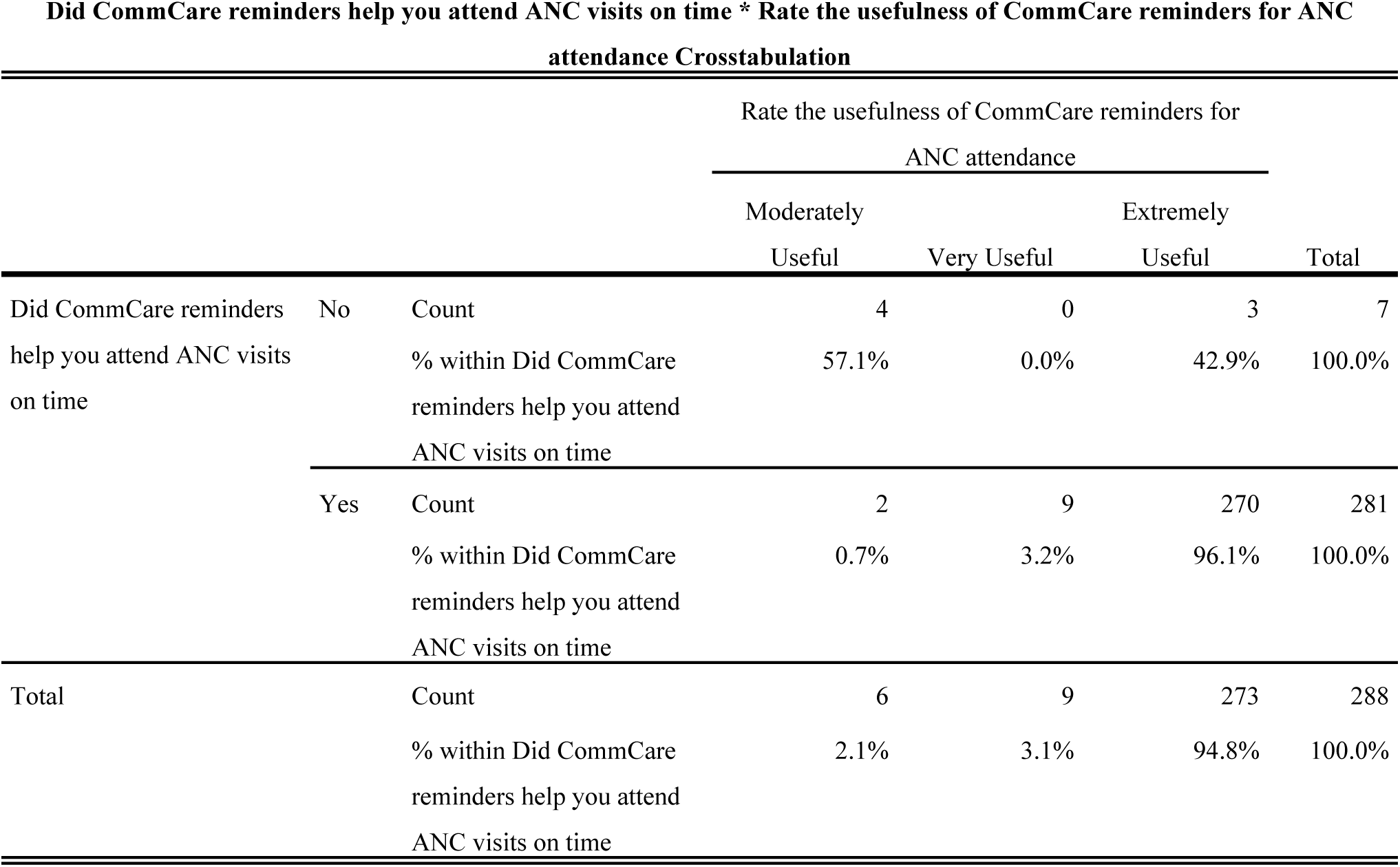
Relationship Between ANC Visit through CommCare Remainders and Usefulness of CommCare Remainders for ANC attendance.

**Table 2.3.** Chi Square tests for the Above.

|  | Value | df | Asymptotic Significance (2-sided) |
| --- | --- | --- | --- |
| Pearson Chi-Square | 106.663<sup>a</sup> | 2 | .000 |
| Likelihood Ratio | 25.197 | 2 | .000 |
| N of Valid Cases | 288 |  |  |

a. 2 cells (33.3%) have expected count less than 5. The minimum expected count is .15.

### PNC Follow-up and Perceived Care Quality

PNC follow-up within first week postpartum rose 40%→80% (χ²=6.12, *p*=0.014). Overall, 96.2% reported that reminders helped them attend PNC on time; χ²=28.451, *p*<0.001.

Beyond attendance, CommCare also shaped mothers’ confidence in handling newborn care. 93.8% of participants reported increased confidence after CommCare-assisted PNC visits Only a small fraction (6.3%) felt no improvement. CommCare visits made postnatal care more consistent with 95.8% of mothers noted greater regularity compared with past experiences without the platform (Table 3.1-3.2)

**Table 3.1.** Relationship Between CommCare Remainders Usefulness for PNC Follow-up and PNC Attendance Influence by CommCare Remainders.

| Rate the usefulness of CommCare reminders for PNC follow-up * Did CommCare reminders help you attend PNC checkups on time Crosstabulation |  |  |  |  |  |
| --- | --- | --- | --- | --- | --- |
|  |  |  | Did CommCare reminders help you attend PNC checkups on time |  | Total |
|  |  |  | No | Yes |  |
| Rate the usefulness of CommCare reminders for PNC follow-up | 2 | Count | 0 | 1 | 1 |
|  |  | % within Rate the usefulness of CommCare reminders for PNC follow-up | 0.0% | 100.0% | 100.0% |
|  | 3 | Count | 3 | 7 | 10 |
|  |  | % within Rate the usefulness of CommCare reminders for PNC follow-up | 30.0% | 70.0% | 100.0% |
|  | 4 | Count | 0 | 1 | 1 |
|  |  | % within Rate the usefulness of CommCare reminders for PNC follow-up | 0.0% | 100.0% | 100.0% |
|  | 5 | Count | 5 | 271 | 276 |
|  |  | % within Rate the usefulness of CommCare reminders for PNC follow-up | 1.8% | 98.2% | 100.0% |
| Total |  | Count | 8 | 280 | 288 |
|  |  | % within Rate the usefulness of CommCare reminders for PNC follow-up | 2.8% | 97.2% | 100.0% |

**Table 3.2.**
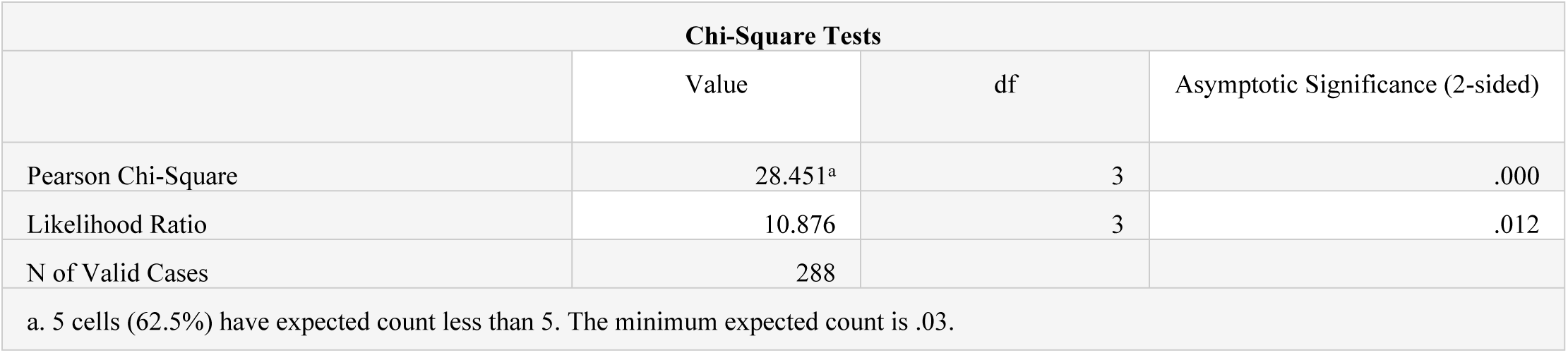
Chi Square Tests of the Above.

| Chi-Square Tests |  |  |  |
| --- | --- | --- | --- |
|  | Value | df | Asymptotic Significance (2-sided) |
| Pearson Chi-Square | 28.451<sup>a</sup> | 3 | .000 |
| Likelihood Ratio | 10.876 | 3 | .012 |
| N of Valid Cases | 288 |  |  |
| a. 5 cells (62.5%) have expected count less than 5. The minimum expected count is .03. |  |  |  |

#### 3.4 Immunization Uptake and Adherence

Immunization adherence increased 60%→82% (χ²=11.34, *p*=0.002); missed appointments fell by 24%. χ²=55.524, *p*<0.001 signaled fewer missed immunization visits among those receiving reminders (Φ=−0.439; Cramér’s V=0.439; contingency coefficient=0.402). Despite gains, 34.4% of children under two were not fully immunized, pointing to persistent access barriers (transport, socio-economic constraints, and hesitancy) (Table 4.1-4.3)

**Table 4.1:** Caregivers’ Perceptions of CommCare Reminders and Child Immunization Outcomes.

|  |  | Count | Table N % | 95.0% Lower CL<br>for Table N % | 95.0% Upper CL<br>for Table N % |
| --- | --- | --- | --- | --- | --- |
| Did CommCare reminders influence your decision to immunize your child | No | 10 | 3.5% | 1.8% | 6.1% |
|  | Yes | 278 | 96.5% | 93.9% | 98.2% |
| Is your child under 2 years fully immunized for their age | No | 99 | 34.4% | 29.1% | 40.0% |
|  | Yes | 189 | 65.6% | 60.0% | 70.9% |
| Did you receive timely reminders for immunization schedules through CommCare | No | 18 | 6.3% | 3.9% | 9.5% |
|  | Yes | 270 | 93.8% | 90.5% | 96.1% |
| Were there fewer missed immunization appointments after CommCare was introduced | No | 214 | 74.3% | 69.0% | 79.1% |
|  | Yes | 74 | 25.7% | 20.9% | 31.0% |
| Would you recommend CommCare-assisted CHW immunization services to other caregivers | Yes | 288 | 100.0% | . | . |

**Table 4.2.** Relationship of Immunization Remainders through CommCare and Immunization scheduled missed.

|  |  | Were there fewer missed immunization appointments after CommCare was introduced |  | Total |
| --- | --- | --- | --- | --- |
|  |  | No | Yes |  |
| Did you receive timely reminders for immunization schedules through CommCare | No | 0 | 18 | 18 |
|  | Yes | 214 | 56 | 270 |
| Total |  | 214 | 74 | 288 |

**Table 4.3.** Chi Square Test of the Above.

|  | Value | df | Asymptotic Significance (2-sided) | Exact Sig. (2-sided) | Exact Sig. (1-sided) |
| --- | --- | --- | --- | --- | --- |
| Pearson Chi-Square | 55.524<sup>a</sup> | 1 | .000 | .000 | .000 |
| Continuity Correction<sup>b</sup> | 51.451 | 1 | .000 |  |  |
| Likelihood Ratio | 52.555 | 1 | .000 | .000 | .000 |
| Fisher's Exact Test |  |  |  | .000 | .000 |
| N of Valid Cases | 288 |  |  |  |  |

a. 1 cells (25.0%) have expected count less than 5. The minimum expected count is 4.63.

#### 3.5 Additional analysis — Program reach

Statistical analysis revealed that phone ownership did not significantly predict exposure to CommCare reminders (χ² = 0.657, df = 1, p = 0.418). Fisher’s exact test, often more reliable in small subgroups, confirmed the absence of a meaningful association (p = 0.476). The effect size was negligible (Φ = −0.048; Cramér’s V = 0.048), indicating that the relationship between owning a phone and being reached by CommCare was essentially nonexistent.

To further test robustness, a binary logistic regression was performed with CommCare exposure as the dependent variable and phone ownership as the independent predictor. The model explained very little variance (Nagelkerke R² = 0.003), and the odds ratio for phone owners being exposed to CommCare was 0.92 (95% CI: 0.48–1.76, p = 0.814), confirming that phone access neither increased nor decreased the likelihood of benefiting from the intervention.

|  | Value | df | Asymptotic<br>Significance<br>(2- sided) | Exact<br>Sig.<br>(2- sided) | Exact<br>Sig.<br>(1- sided) |
| --- | --- | --- | --- | --- | --- |
| Pearson Chi-Square | .657<sup>a</sup> | 1 | .418 |  |  |
| Continuity Correction<sup>b</sup> | .397 | 1 | .529 |  |  |
| Likelihood Ratio | .681 | 1 | .409 |  |  |
| Fisher's Exact Test |  |  |  | .476 | .269 |
| N of Valid Cases | 288 |  |  |  |  |

a. 0 cells (0.0%) have expected count less than 5. The minimum expected count is 12.25.
b. Computed only for a 2x2 table

## Discussion

This study demonstrated consistent improvements in maternal and child health service uptake through CommCare-assisted reminders, particularly in relation to timeliness and continuity of care. Perceived usefulness was strikingly high: the vast majority of respondents (94.8%) rated reminders as “extremely useful” for antenatal attendance, while only small proportions selected “moderately useful” (2.1%) or “very useful” (3.1%). Among those who reported that reminders helped them attend on time, 96.1% described them as extremely useful, whereas in the small “No” subgroup, perceptions of usefulness skewed toward moderate. These findings underscore the close alignment between perceived utility and timely care-seeking.

### Summary of Main Findings

Patterns of attendance reinforced these perceptions. A crosstabulation of “reminders helped you attend on time” against “ANC visits during last pregnancy” showed a marked shift toward higher visit volumes. All women who reported benefiting from reminders had at least four contacts, and 119 of them reached five or more visits, compared with none in the group that reported no benefit. Early initiation also improved substantially: first-trimester ANC uptake rose from 45% before the intervention to 65% afterward (χ² = 8.34, p = 0.004), while the proportion achieving the recommended four or more ANC visits increased from 55% to 78% (χ² = 7.72, p = 0.009). These findings parallel gains in postnatal care, where follow-up within the first week postpartum doubled from 40% to 80% (χ² = 6.12, p = 0.014). These results are consistent with existing literature. A study in Tanzania by (17) found that SMS-based interventions in Zanzibar increased the proportion of women completing four or more ANC visits and improved perinatal outcomes, effects attributed to reminder-driven timeliness and continuity.

Similarly, a systematic review concluded shows that mobile text message reminders improved both coverage and timeliness of childhood vaccination in low- and middle-income countries (8). What distinguishes the present findings is the delivery pathway. Phone ownership was not significantly associated with CommCare exposure (χ² = 0.657, p = 0.418; Fisher’s p = 0.476; Φ = −0.048), underscoring that the program’s effects were mediated through CHW case management rather than client device ownership. Taken together, the pattern across ANC, PNC, and immunization demonstrates that reminder systems embedded in frontline workflows are not only acceptable but highly effective in improving adherence. However, persistent coverage gaps indicate that reminders alone are insufficient to overcome deeper systemic barriers. These findings suggest that combining mHealth reminders with structural supports such as transport vouchers, defaulter tracing, and targeted community engagement may offer a more comprehensive approach to closing the last mile of maternal and child health service delivery.

### Comparison to Existing Literature

The CommCare intervention produced a dramatic improvement in postnatal follow-up: attendance within the first week postpartum rose from 40% to 80% (χ² = 6.12, p = 0.014). Moreover, 96.2% of respondents reported that reminders helped them attend PNC on time (χ² = 28.451, p < 0.001). Beyond mere attendance, the program also strengthened mothers’ self-efficacy in newborn care: 93.8% of participants stated that their confidence improved after CommCare-supported PNC visits, with only 6.3% reporting no change. In terms of consistency, 95.8% of mothers observed that PNC visits through CommCare were more regular compared to their experiences before the intervention. These findings resonate with broader mHealth evidence on strengthening PNC and maternal-neonatal outcomes. A systematic review of rural mHealth communication interventions found that mobile messaging often enhances maternal self-efficacy and confidence with newborn care through reinforcing attitudes, perceived norms, and practical skills (17,18). The review notes that improved confidence (self-efficacy) is a recurring theme across studies, arising when mothers attend follow-ups and receive supportive education (19).

In Nigeria, a qualitative exploration of mothers’ experiences with postnatal mHealth interventions confirmed that reminder messages provide reassurance and reinforce knowledge about baby care practices, thereby promoting utilization of PNC services (20). In Addis Ababa, enhanced reminders (via SMS or voice calls) at 48 and 24 hours before scheduled PNC appointments significantly increased compliance: women in the intervention arm had nearly triple the odds of adherence (AOR ≈ 2.98, 95% CI 1.51–5.80) compared to controls (p = 0.005) (14). The analysis revealed significant improvements in childhood immunization uptake and continuity following the introduction of CommCare reminders. Adherence increased markedly from 60% at baseline to 82% post-intervention, a difference that was statistically significant (χ² = 11.34, p = 0.002). Correspondingly, the proportion of missed appointments fell by 24%. A strong chi-square signal (χ² = 55.524, p < 0.001) confirmed that reminders were associated with fewer missed immunization visits. Effect size measures further supported this association, with moderate to strong correlations (Φ = −0.439; Cramér’s V = 0.439; contingency coefficient = 0.402), demonstrating that the reminder system substantially influenced caregiver behavior(14).

### Implications for Practice and Policy

The findings of this study underscore the importance of institutionalizing CommCare within routine district health operations rather than treating it as a stand-alone innovation. Embedding the platform into scheduling systems and defaulter tracing mechanisms ensures that reminders are not just sent, but acted upon through systematic follow-up by CHWs. This integration is critical for converting digital prompts into completed visits, especially in resource-limited rural contexts where the gap between intention and actual service uptake remains wide. Provision for CHWs airtime, device maintenance, and charging infrastructure must be considered non-negotiable program inputs. Without reliable access to communication and power, the continuity and timeliness of digital case management break down, eroding both caregiver trust and health worker motivation. The evidence from this study suggests that such logistical supports directly underpin the consistency of PNC follow-up and immunization adherence, which were markedly higher among those reached via CommCare. These dimensions are vital for overcoming persistent barriers such as vaccine hesitancy and gendered decision-making dynamics that limit women’s autonomy in health-seeking. Digital interventions like CommCare must therefore be embedded in a broader communication strategy that resonates with local values while promoting evidence-based practices. By embedding digital case management into daily workflows, providing adequate logistical support, and aligning with community and system realities, policymakers can ensure that the gains in ANC, PNC, and immunization adherence observed in this study are not only sustained but scaled across similar low-resource settings.

### Strengths and Limitations

This study has several notable strengths. First, it was implemented in a real-world, program-embedded context within a hard-to-reach district, ensuring that the results reflect the practical realities of routine service delivery rather than idealized pilot conditions. The deployment was not an isolated research exercise but integrated into district health structures, which enhances both the ecological validity and the likelihood of scalability. This study drew on a sufficiently large sample size (n = 288), providing adequate statistical power to detect meaningful associations across antenatal care (ANC), postnatal care (PNC), and immunization outcomes. The convergence of evidence across multiple service indicators timeliness of ANC initiation, early PNC follow-up, and immunization adherence strengthens the internal consistency of the findings.

Nevertheless, important limitations should be acknowledged. The cross-sectional design restricts the ability to make causal claims about the observed associations. While chi-square tests and effect sizes demonstrated strong relationships, temporality and confounding factors cannot be definitively ruled out without longitudinal or experimental designs. Another limitation is the reliance on self-reported measures for some indicators, which may introduce recall bias or social desirability effects, particularly in responses related to confidence in newborn care. The absence of complementary qualitative data from caregivers and CHWs further constrains the interpretation of why reminders worked effectively in some cases but less so in others. Finally, there is the potential for selection bias, as individuals who were more likely to attend services may also have been more likely to be exposed to and benefit from CommCare, thereby inflating observed effects.

### Future Research

Future research should move beyond single-site, cross-sectional evaluations to multi-district and multi-facility studies that capture a broader spectrum of maternal and child health service environments. Employing longitudinal or quasi-experimental designs would allow researchers to more rigorously assess causal relationships and track the sustained impact of CommCare over time on adherence to antenatal, postnatal, and immunization schedules. In addition, there is a need for mixed-methods approaches that complement quantitative outcome data with qualitative insights from caregivers, CHWs, and facility staff. Exploring the equity dimensions of CommCare implementation should also be a priority. While this study found that phone ownership did not predict exposure, further work could examine how socioeconomic status, literacy, and geographic remoteness interact with CHW-mediated digital tools. Finally, future studies should investigate the system-level effects of embedding CommCare into district workflows, particularly its influence on staff workload, supply chain coordination (e.g., vaccine availability), and the quality of patient-provider interactions

## Methods

### Study design and setting

Facility-based, cross-sectional quantitative design at Falaba CHC (BeMOC) and its catchment communities in Sulima Chiefdom was done in August-September 2025, a mountainous, remote area with seasonal access constraints. Services include ANC, PNC, immunization, malaria/HIV care, and nutrition. CommCare operates through MoHS/DHMT-INGO partnerships. The tool has been in use since the start of this World Vision Financial Year FY25. Before the advent CHWs were using the traditional paper-based method.

### Population and sample

The study population consisted of women of reproductive age (including pregnant/lactating) and caregivers of children 0–59 months. The sample size for this study was calculated using Fisher’s formula. Where the PHU Catchment population for the above category is n=6868. And using an assumption of 95% confidence interval (Z = 1.96), an estimated proportion of 50% (p = 0.5), and a margin of error of ±12.3% (E = 0.123), the required minimum sample size N=288 participated via primary data collection with CommCare-aligned questionnaires.

### Instrument and variables

A structured CommCare tool covered Section A: demographics; B: ANC; C: PNC; D: immunization; E: perceptions/recommendations. Outcomes included: (i) *timely ANC* (first-trimester initiation) and ≥4 visits; (ii) *PNC follow-up* (any visit and first-week postpartum visit); (iii) *immunization adherence* and missed appointments. The instruments were tested at the Gberia Timbakor CHP among study participants with similar demographic characteristics.

### Data management and analysis

Data was downloaded from the CommCare server, exported to excel for cleaning and was analyzed using IBM SPSS version 25. Descriptive statistics were summarized with frequencies/percentages. Chi-square (χ²) tests of association were conducted to assess associations and relationships for 2×2 tables, Fisher’s Exact was used as needed; symmetric measures (Φ, Cramér’s V) quantified effects. Significance at α=0.05.

### Ethics

Ethical approval was obtained from the Njala University Institutional Review Board (Reference: NU/IRB/PH/2024/034). Permission to conduct the study was granted by the District Health Management Team (DHMT) Falaba, chiefdom stakeholders and the management of the Falaba CHC. Written informed consent was obtained from all respondents prior to data collection, and confidentiality and anonymity were maintained throughout the study (including encryption of CommCare data).

## Data Availability

The data of this manuscript is available with me and will be posted if need. The data has been extracted from CommCare Hq.

## ACKNOWLEDGEMENTS

The authors would like to express their sincere gratitude to the District Health Management Team (DHMT) of Falaba and the management of Falaba Community Health Center for their cooperation and support throughout the study. We also extend our appreciation to the community stakeholders and participants; whose time and openness made this research possible. Special thanks go to the community health workers for their commitment in implementing CommCare and assisting with data collection. We acknowledge the academic guidance and technical review provided by faculty members at the School of Public Health, Njala University. Finally, we thank World Vision Sierra Leone and partnering organizations for their operational collaboration in the deployment of CommCare in the study district.

## SUPPORTING INFORMATION CAPTIONS

**Table 5: Demographic Characteristics of Respondent**

**Table 6.1 CommCare Remainders and ANC visits**

**Table 2.2 Chi-Square Test of CommCare Remainders and ANC Visit**

**Table 2.3 Relationship Between ANC Visit through CommCare Remainders and Usefulness of CommCare Remainders for ANC attendance.**

**Table 2.4 Chi Square tests for the Above**

**Table 7 Relationship Between CommCare Remainders Usefulness for PNC Follow-up and PNC Attendance Influence by CommCare Remainders.**

**Table 3.1 Chi Square Tests of the Above**

**Table 8: Caregivers’ Perceptions of CommCare Reminders and Child Immunization Outcomes**

**Table 4.1 Relationship of Immunization Remainders through CommCare and Immunization scheduled missed.**

**Table 4.2 Chi Square Test of the Above**

